# Increasing Heroin, Cocaine, and Buprenorphine Arrests Reported to the Maine Diversion Alert Program

**DOI:** 10.1101/19003376

**Authors:** Kevin J Simpson, Matthew T Moran, Kenneth L McCall, John Herbert, Michelle Foster, Olapeju M Simoyan, Dipam T Shah, Clare Desrosiers, Stephanie D Nichols, Brian J Piper

**Affiliations:** Geisinger Commonwealth School of Medicine, Scranton, PA 18509, USA; University of New England, Portland, ME 04103, USA; Diversion Alert, Houlton, ME 04730, USA; Geisinger Marworth Treatment Center, Waverly, PA 18471; Husson University School of Pharmacy, Bangor, ME 04401, USA; Tufts University, Medford, MA 02155, USA; Center for Pharmacy Innovations and Outcomes, Forty Fort, PA 18704 USA

## Abstract

**Background:** The opioid overdose crisis is especially pronounced in Maine. The Diversion Alert Program (DAP) was developed to combat illicit drug use and prescription drug diversion by facilitating communication between law enforcement and healthcare providers with the goal of limiting drug-related harms and criminal behaviors. Our objectives in this report were to analyze 2014-2017 DAP for: 1) trends in drug arrests and, 2) differences in arrests by offense, demographics (sex and age) and by region.

**Methods:** Drug charges (N = 8,193, 31.3% female, age = 33.1 ± 9.9) reported to the DAP were examined by year, demographics, and location.

**Results:** The most common substances of the 10,064 unique arrests reported were heroin (N = 2,203, 21.9%), crack/cocaine (N = 945, 16.8%), buprenorphine (N = 812, 8.1%), and oxycodone (N = 747, 7.4%). While the overall number of arrests reported to the DAP declined in 2017, the proportion of arrests involving opioids (heroin, buprenorphine, or fentanyl) and stimulants (cocaine/crack cocaine, or methamphetamine), increased (*p* < .05). Women had significantly increased involvement in arrests involving sedatives and miscellaneous pharmaceuticals (e.g. gabapentin) while men had an elevation in stimulant arrests. Heroin accounted for a lower percentage of arrests among individuals age ≥ 60 (6.6%) relative to young-adults (18-29, 22.3%, *p* < .0001). Older-adults had significantly more arrests than younger-adults for oxycodone, hydrocodone, and marijuana.

**Conclusion:** Heroin had the most arrests from 2014-2017. Buprenorphine, fentanyl and crack/cocaine arrests increased appreciably suggesting that improved treatment is needed to prevent further nonmedical use and overdoses. The Diversion Alert Program provided a unique data source for research, a harm-reduction tool for health care providers, and an informational resource for law enforcement.

## Introduction

Despite ongoing reductions in opioid prescribing [1,2], deaths in the United States from opioid overdoses continue to climb [3]. The contrast between a reduced supply of licit opioids and a further escalation of opioid deaths is especially evident in Maine. Opioid prescribing volume in the US declined by 11% from 2016 to 2017 while Maine had the largest decline of nearly 25% [4]. Maine experienced an 11.2% increase in overdose deaths from 2016 to 2017. The profile of decedents was predominately (71.5%) male, middle-aged, with four-fifths (78.9%) testing positive for multiple drugs [5]. Further, Maine had the second highest rate in the US of pregnant women with an opioid use disorder (34.1 / 1,000) [6]. It is unclear whether the epidemic is being driven by increased opioid demand or supply, given that rates of deaths of despair (suicide and unintentional overdose) involving opioids have increased 2.5 fold (17% to 42%) since 2000, suggesting increased demand [7]. Although opioids continue to receive the most attention, there have also been concerns about increased overdoses involving cocaine [3].

Maine has adopted several new public health policies and programs in an attempt to fetter and ultimately reverse this deadly trend. Recent laws have mandated use of the Prescription Drug Monitoring Program (PDMP) by prescribers and pharmacists, implemented dose and supply limits for opioid prescriptions and increased access to naloxone [1, 8, 9]. Furthermore, strategies such as the Diversion Alert Program (DAP) provided information to medical professions regarding persons arrested for offenses involving prescription or recreational drugs [10, 11]. The DAP was operational from 2013 until 2018 and provided a unique state-level resource which aimed to reduce prescription drug diversion. Law enforcement reported arrests involving prescription or illicit drugs to a database accessible to prescribers and pharmacists. The DAP has been reported to increase communication between healthcare providers and patients with suspected nonmedical drug use behaviors [12]. The DAP has also demonstrated utility as a public health tool to evaluate patterns in drug usage [5, 10, 11, 13, 14].

Many criminal justice systems make a distinction between possession and trafficking. The intentional or knowing possessing of what is known or believed to be a scheduled drug is possession. There are minimum thresholds like 200 mg of heroin, fentanyl, oxycodone, hydrocodone, or methamphetamine or 2 g of cocaine to classify as possession. Trafficking may refer to creating, manufacturing or being arrested with larger weights (e.g. 2 g or > 90 containers of heroin or fentanyl powder; 14 g methamphetamine or cocaine; 4 g cocaine base, 800 mg of oxycodone, 200 mg of hydromorphone) of a controlled substance [15]. Over three-quarters of persons reported to the DAP in 2014 for trafficking controlled prescription medications did not have a record for this substance in the state’s Prescription Monitoring Program [10] indicating that these databases provide complementary, but non-redundant, information.

Buprenorphine availability and use in Maine [5] parallels the national pattern [1] with appreciable gains [5]. Almost half of opioid prescriptions to young-adults in their twenties was for buprenorphine [13]. However, of the top five most prescribed drugs based on Maine insurance claims, more money was spent on the buprenorphine and naloxone film (Suboxone, $16.8 million) in 2017 than on the other four agents (hydrochlorthiazide, ProAir, Ventolin, and omeprazole) combined ($12.7 million) [16]. Buprenorphine, when dosed flexibly, and methadone equally suppressed illicit opioid use but buprenorphine was inferior to methadone in treatment retention. Fixed buprenorphine doses of 16 mg/day or greater were no different in efficacy from methadone [17]. The first-line of the warnings and precautions section of the US package insert for the film notes “buprenorphine can be abused in a similar manner to other opioids”. This is key information as there is an increasing evidence about nonmedical buprenorphine use including diversion and injection [18-20]. The US street value per mg of buprenorphine was over double that of methadone, oxycodone, and hydrocodone and four-fold that of morphine [21]. Importantly, nonmedical use of buprenorphine is safer than nonmedical use of full opioid agonists via a reduction in overdose risk. In the general population, methadone was associated with over six-times more fatal overdoses than buprenorphine.

Our objectives in this report were to analyze the 2014-2017 Maine DAP data for trends in arrests for both illicit (e.g. heroin) and prescription drugs (e.g. buprenorphine) including by year, region, arrestee demographic (sex and age), and offense (possession versus trafficking).

## Materials and Methods

### Subjects

Subjects included all individuals age 18 or older reported to the Maine DAP for drug crimes (N = 8,193, 31.27% female, mean age = 33.07, SD = 9.92, Min = 18, Max = 83). Information regarding sex, age, offense, drug(s), arrest date, town of residence, and reporting agency were submitted by city, county, state, and federal law enforcement personnel.

### Procedures

Substances were categorized into drug classes: 1) opioids (e.g. heroin, methadone); 2) stimulants (e.g. cocaine, amphetamines); 3) sedatives (e.g. benzodiazepines, barbiturates, zolpidem); 4) hallucinogens / other (e.g. psychedelic mushrooms, marijuana, alpha-pyrrolidinopentiophenone (a-PVP), methylenedioxymethamphetamine (MDMA)); 5) miscellaneous pharmaceutical including substances not included in 1-4 that are typically manufactured by a pharmaceutical company (e.g. gabapentin); and 6) other (e.g. unspecified drugs, inhalants, drug paraphernalia). These classes have been used previously in single-year reports [5, 14]. Drugs were further classified in accordance to the US 1970 Controlled Substances Act as Schedule I-V, non-controlled prescription, or over the counter (e.g. pseudoephedrine). Arrestee age was categorized into six groups, 18-29 (41.07%), 30-39 (34.46%), 40-49 (13.85%), 50-59 (6.19%), ≥ 60 (1.43%), and age not reported (3.00%). A regional analysis was completed by examining the county of residence of the arrestee. The

Institutional Review Board approved this study and procedures were in accordance with the 2004 revision of the Helsinki Declaration.

### Data-analysis

The DAP began in June, 2013 and ceased operation due to unstable funding in early 2018. Only full-years (2014 – 2017) were examined. The term arrests will be used interchangeably with all other types of drug charges reported including summons or indictments for this report. Arrest offenses were categorized into possession, furnishing, possession with intent to distribute, trafficking, manufacturing, operating under the influence (OUI), and other. For examination of arrests involving multiple drugs (Max = 12), each unique drug reported per offense was the unit of analysis [11, 14]. Epi Info, version 7.2.1.0 was used to analyze the data. Figures were created using GraphPad Prism, version 7.04. Fisher’s Exact Test or chi-square test with Yates’ Correction for non-parametric analyses were performed using GraphPad chi-square calculator. Unpaired t-tests were performed using GraphPad t-test calculator. An alpha of < .05 was considered significant although statistics that met more conservative thresholds were noted. Heat maps were created with CARTO. County and state populations were derived from the US Census Bureau population estimates for April 1 of each year analyzed. The Arrest Ratio (AR) was calculated as the percent arrests (number arrests countywide / total arrests statewide) divided by the percent population (county population / Maine total population). An AR >1 indicate more arrests in a county than expected based on population. Variability was expressed as the standard deviation (SD).

## Results

Drug arrests were reported to the DAP by local (52.57%), county (7.99%), state (33.30%), and federal (6.14%) agencies. After accounting for arrests involving multiple drugs, there were 10,064 unique charges. Of the 1,353 arrestees with multiple charges, over two-thirds were male (68.37%) or under forty (73.02%).

Figure 1A shows the percentage of charges for each drug class by year and tested for changes relative to 2014. Opioids accounted for over two-fifths of arrests which remained stable with the exception of an increase in 2016 (*p* ≤ .0003). Stimulants were the second most common class with elevations in 2016 (*p* < .01) and 2017(*p* <0.01). The miscellaneous pharmaceuticals class accounted for less than one-tenth of arrests and was transiently increased in 2015 (*p* ≤ .0008). Hallucinogens and the other classes were each significantly (*p* ≤ .01) lower each year after 2014. Figure 1B further examines some of the most common drugs by year. Heroin, buprenorphine, alprazolam, methamphetamine, and fentanyl were significantly increased in 2017 relative to 2014 while unspecified agents and oxycodone decreased. Arrests for cocaine and crack cocaine were more frequent than heroin in 2017 (Supplemental Tables 1 and 2).

**Figure 1.**
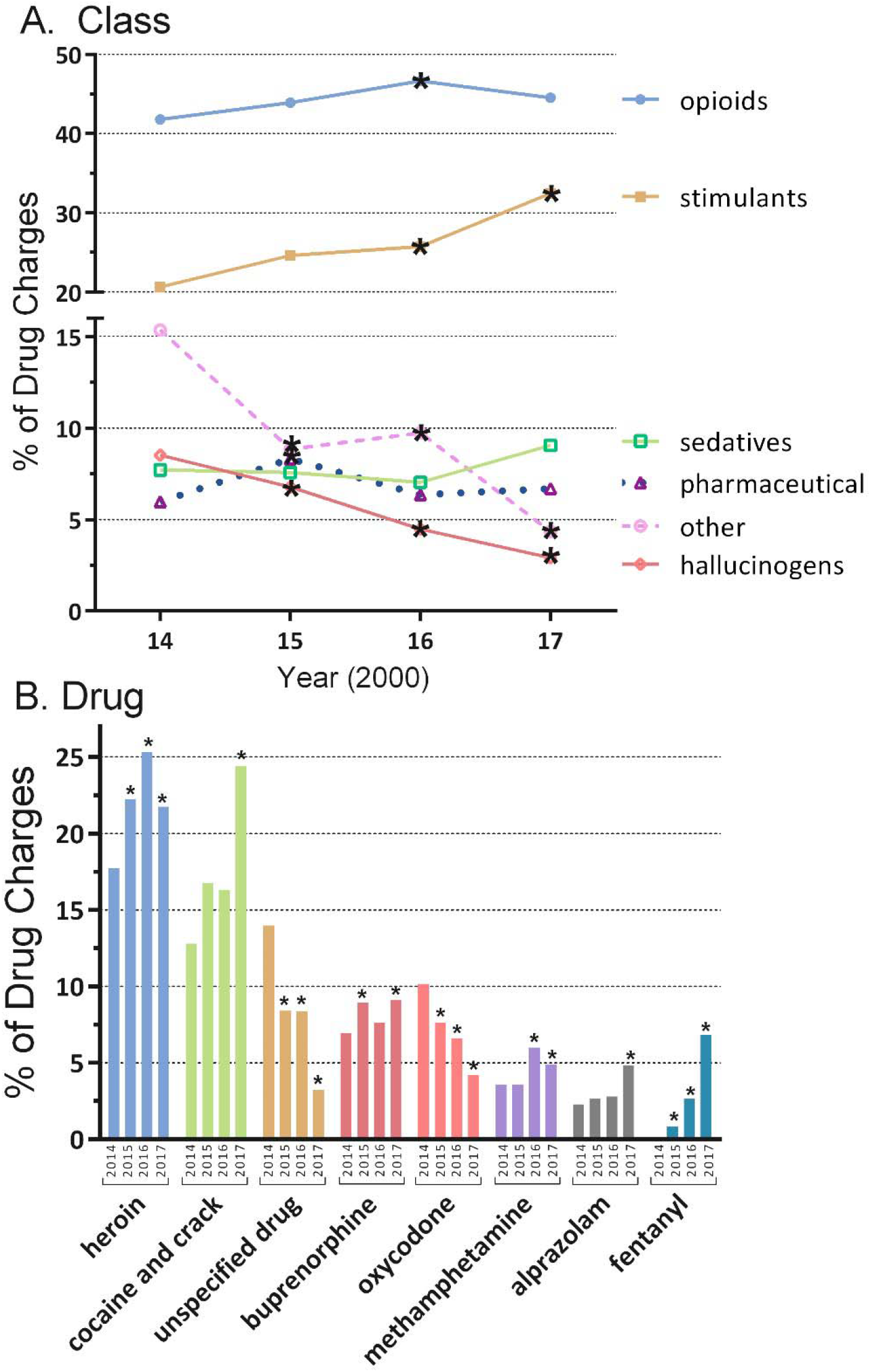
Percent of drug classes (A) and selected drugs (B) by year reported to the Maine Diversion Alert Program. \**p* ≤ .05 versus 2014.

Figure 2 depicts arrests by charge and reveals that, overall, possession accounted for three-fifths (60.0%) of arrests, trafficking one-quarter (24.5%), followed by distribution (3.9%) and possession with intent to distribute (3.5%). The benzodiazepines were most common (≥ 85.5%) for possession. Crack had the highest percentage trafficking (43.7%). One-fifth of fentanyl arrests were for distribution (20.3%) which was twice as common as the next highest drug (cocaine = 9.5%). Manufacturing was fifteen-fold higher for methamphetamine (13.3%) relative to the next most common agent (marijuana = 0.9%). Marijuana led for OUI (6.4%) and other (15.3%) arrests (Supplemental Table 3).

**Figure 2.**
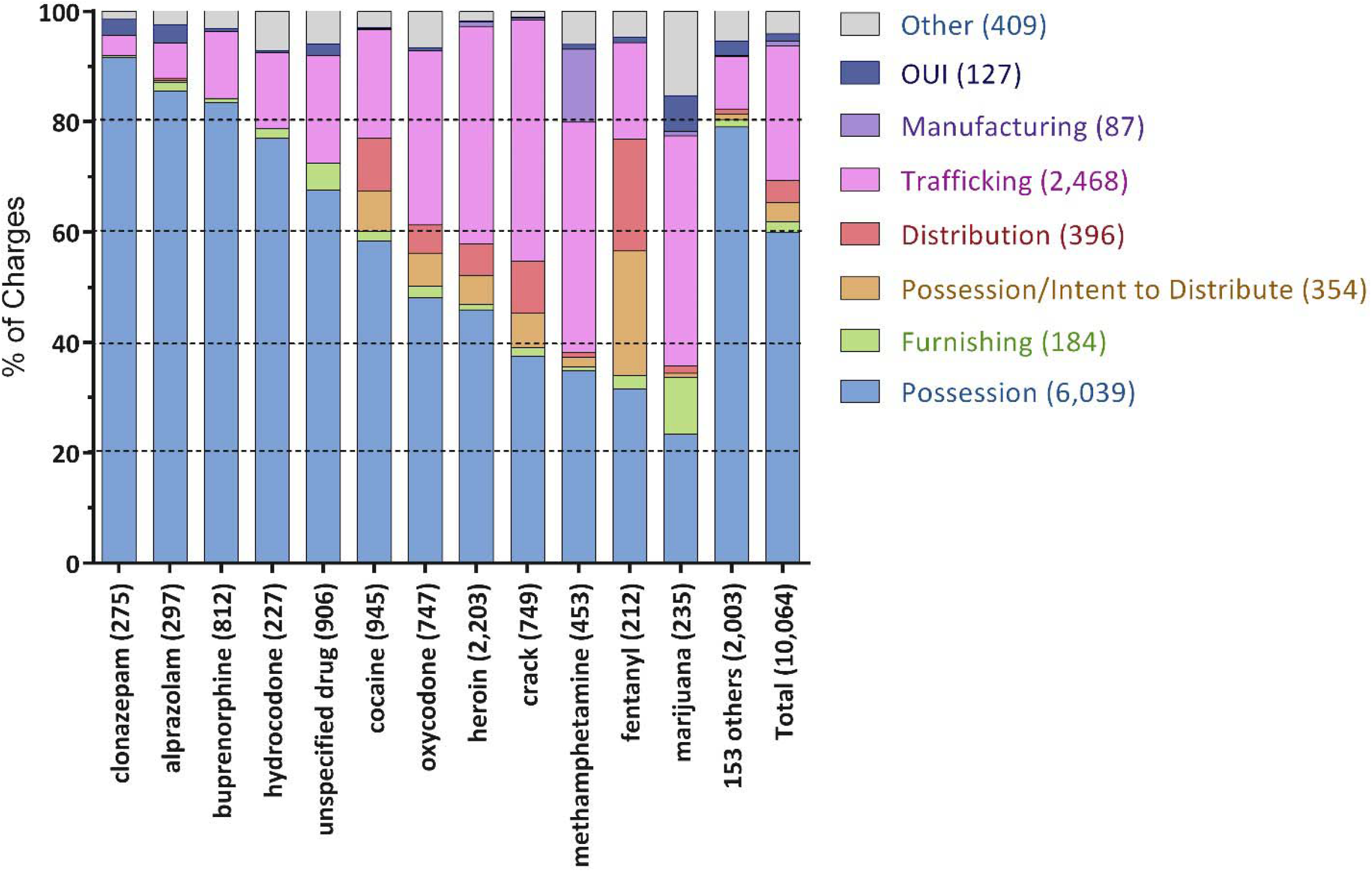
Arrest charge by drug, ranked by percent Possession, reported to the Maine Diversion Alert Program from 2014 to 2017. Operating Under the Influence: OUI. Number of arrests is in parentheses.

The sexes showed broad similarities in the classes and DEA Schedule. However, small, but significant, differences were noted with males being arrested more commonly for stimulants and hallucinogens and females for sedatives and miscellaneous pharmaceutical agents (Figure 3A). Figure 3B depicts the Schedule of drugs (note that Schedule V (N = 12) was not shown due to limited data, see also Supplemental Tables 4-5). Males were more likely to be arrested for Schedule I agents and females for Schedule IV and non-controlled prescriptions. Males (33.29 ± 10.25, Min = 18, Max = 83) and females (32.59 ± 9.16, Min = 18, Max = 67) were equivalent in age.

**Figure 3.**
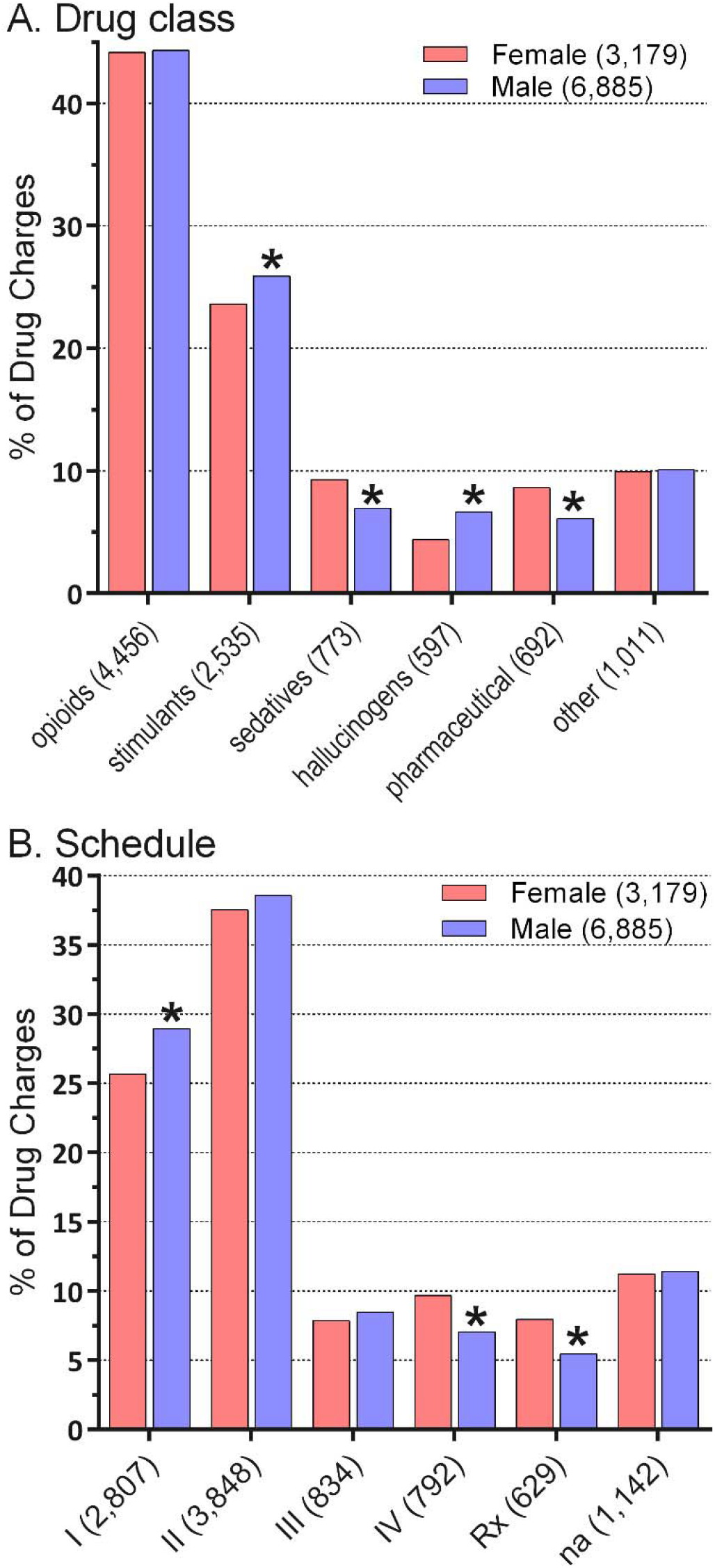
Drug class (A) and schedule (B) by sex reported to the Maine Diversion Alert Program from 2014-2017. Number of arrests are in parentheses. Rx: non-controlled prescription drug.

Figure 4 reports on age differences by drug. Heroin accounted for three-fold more arrests for young-adults (age 18 to 29) than for older-adults (age ≥ 60). Conversely, older-adults were significantly more likely than young-adults to be arrested for oxycodone, hydrocodone, or marijuana (Supplemental Tables 6-8).

**Figure 4.**
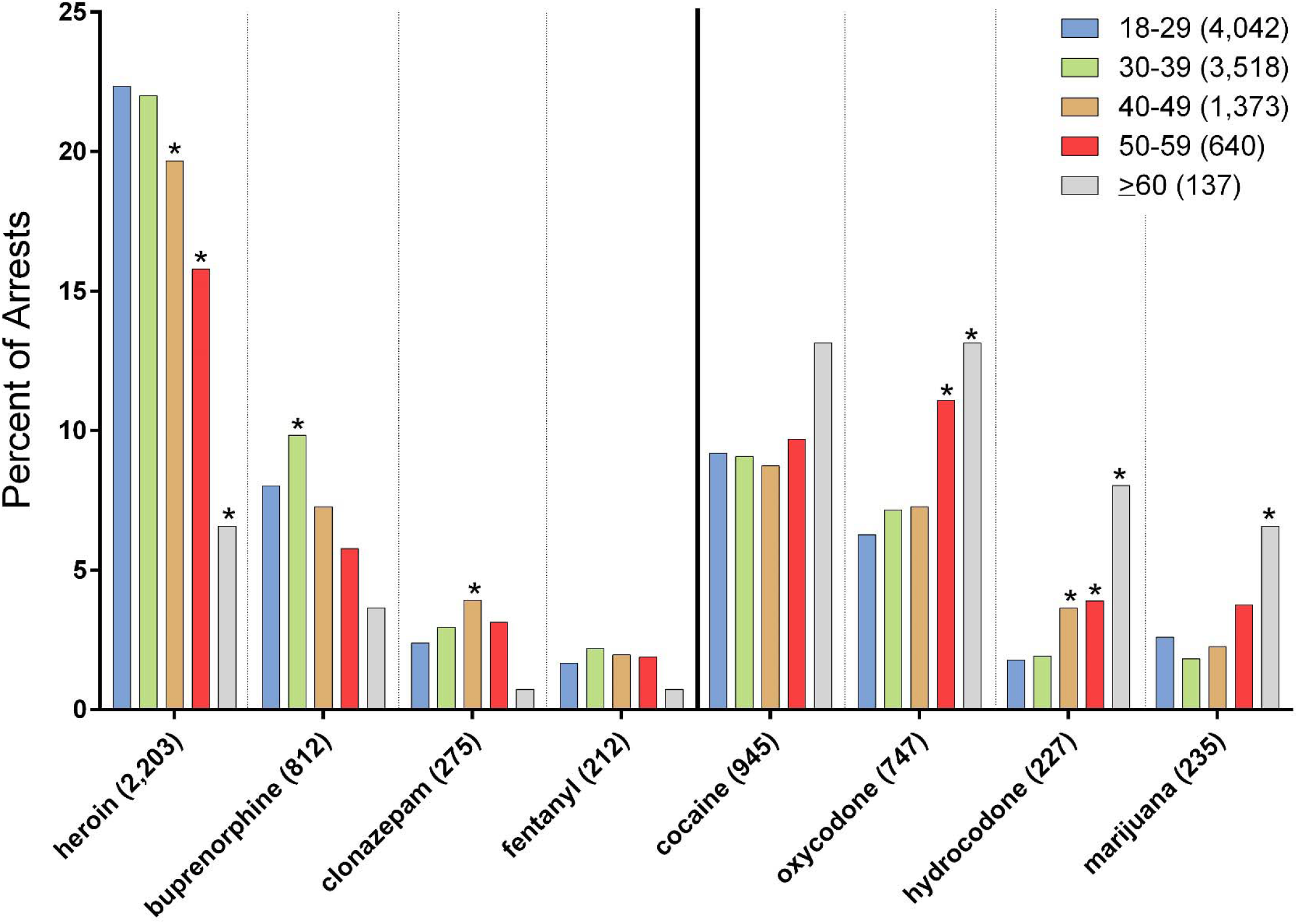
Age differences in drugs reported to the Maine Diversion Alert Program from 2014 to 2017. \**p* < .05 versus 18 – 29.

Arrest data was examined by county. The most arrests occurred in Cumberland (27.15%), Penobscot (12.22%), and Androscoggin (11.32%) which rank first, third, and fifth, respectively, among the sixteen Maine counties for population. Additional information about changes in arrests over time may be found in Supplemental Table 9. A population corrected AR was obtained separately for opioids and stimulants (Supplemental Figure 1, Supplemental Tables 10 and 11). The western and northern counties of Oxford, Franklin, and Piscataquis ranked in the lower third (AR ≤ .70). Only Androscoggin ranked in the upper-third (AR ≥ 1.30) for both opioid and stimulant arrests. The correlation between the AR for opioids and stimulants was not significant (*r*(14) = 0.42, *p* = .11).

**Supplemental Figure 1.**
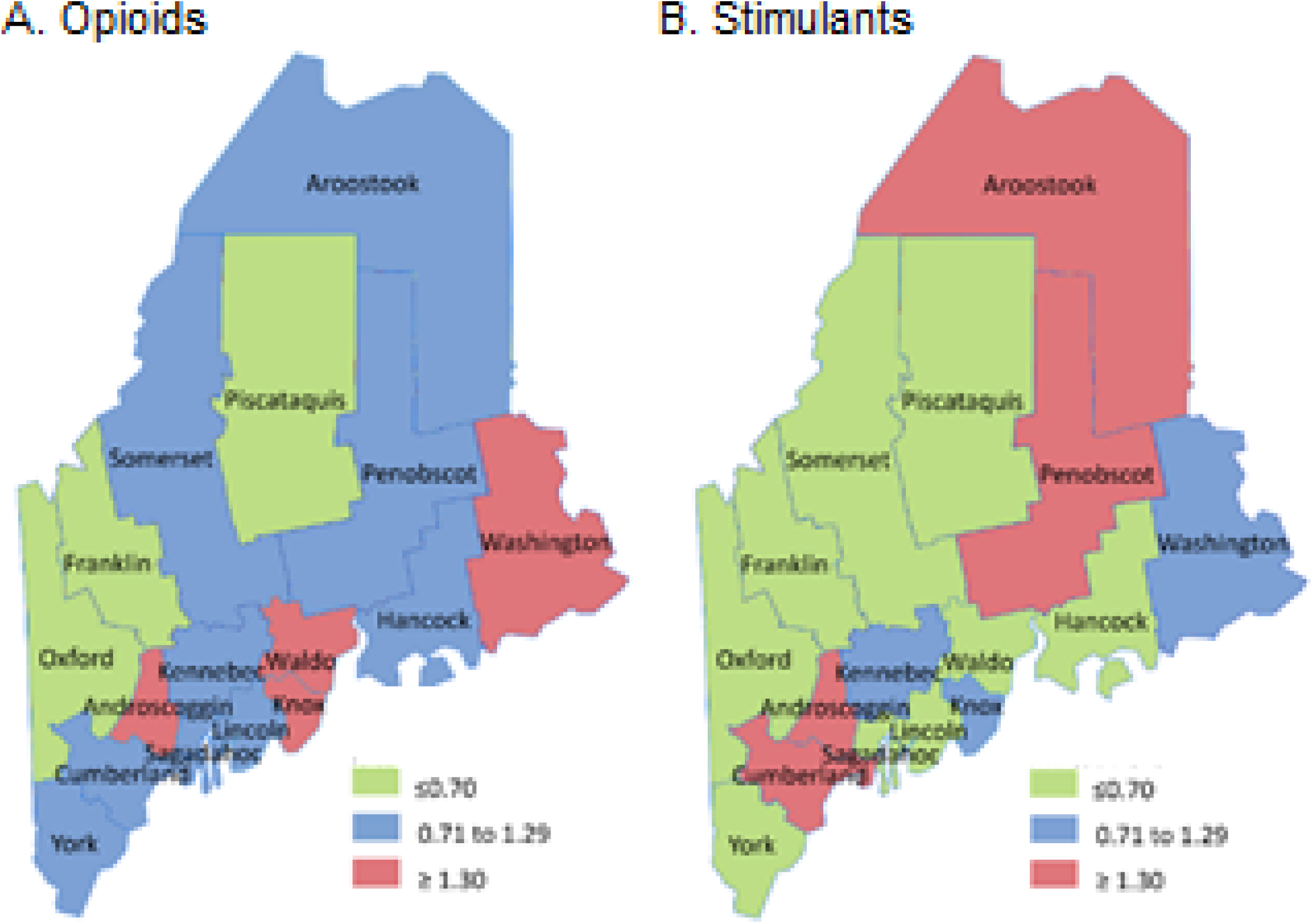
Heat map of the arrest ratio (arrests / population) for opioids (A) and stimulants (B) reported to the Maine Diversion Alert Program from 2014 to 2017.

**Supplemental Table 1.** Frequency of arrests reported to the Maine Diversion Alert Program from 2014-2017 by drug class. ^a^Fisher’s Exact Test *p* values relative to 2014.

| Class |  | Year |  |  |  |  |
| --- | --- | --- | --- | --- | --- | --- |
|  |  | 2014-2017 | 2014 | 2015 | 2016 | 2017 |
| opioids | n | 4,456 | 1,079 | 1,290 | 1,368 | 719 |
|  | % | 44.28% | 41.81% | 43.91% | 46.67% | 44.55% |
|  | *p*<sup>a</sup> | - | - | 0.12 | 0.0003* | 0.08 |
| stimulants | n | 2,535 | 533 | 723 | 755 | 524 |
|  | % | 25.19% | 20.65% | 24.61% | 25.76% | 32.47% |
|  | *p*<sup>a</sup> | - | - | 0.14 | <0.001* | <0.001* |
| sedatives | n | 773 | 199 | 222 | 206 | 146 |
|  | % | 7.68% | 7.71% | 7.56% | 7.03% | 9.05% |
|  | *p*<sup>a</sup> | - | - | 0.84 | 0.35 | 0.13 |
| hallucinogens /<br>other | n | 597 | 220 | 199 | 131 | 47 |
|  | % | 5.93% | 8.52% | 6.77% | 4.47% | 2.91% |
|  | *p*<sup>a</sup> | - | - | 0.01* | <0.001* | <0.001* |
| miscellaneous<br>pharmaceutical | n | 692 | 154 | 244 | 186 | 108 |
|  | % | 6.88% | 5.97% | 8.30% | 6.35% | 6.69% |
|  | *p*<sup>a</sup> | - | - | 0.0008* | 0.58 | 0.36 |
| other | n | 1,011 | 396 | 260 | 285 | 70 |
|  | % | 10.05% | 15.34% | 8.85% | 9.72% | 4.34% |
|  | *p*<sup>a</sup> | - | - | <0.001* | <0.001* | <0.001* |
| Total | n | 10,064 | 2,581 | 2,938 | 2,931 | 1,614 |
|  | % | 100.00% | 100.00% | 100.00% | 100.00% | 100.00% |

**Supplemental Table 2.** Ranking of the most commonly reported drugs to the Maine Diversion Alert Program from 2014-2017. alpha-pyrrolidinopentiophenone: α-PVP. ^a^Fisher’s Exact Test *p* relative to 2014.

| Drug | Year |  |  |  |  |  |  |  |  |  |  |  |
| --- | --- | --- | --- | --- | --- | --- | --- | --- | --- | --- | --- | --- |
|  | 2014 (2,581) |  |  | 2015 (2,938) |  |  | 2016 (2,931) |  |  | 2017 (1,614) |  |  |
|  | n | % | *p* | n | % | *p*<sup>a</sup> | n | % | *p*<sup>a</sup> | n | % | *p*<sup>a</sup> |
| 1. heroin (2,203) | 457 | 17.71% | - | 653 | 22.23% | <0.01* | 742 | 25.32% | <0.01* | 351 | 21.75% | 0.002* |
| 2. cocaine (945) | 178 | 6.90% | - | 314 | 10.69% | <0.01* | 250 | 8.53% | 0.03* | 203 | 12.58% | <0.01* |
| 3. unspecified drug (906) | 361 | 13.99% | - | 247 | 8.41% | <0.001* | 246 | 8.39% | <0.01* | 52 | 3.22% | <0.001* |
| 4. buprenorphine (812) | 179 | 6.94% | - | 263 | 8.95% | 0.006* | 223 | 7.61% | 0.35 | 147 | 9.11% | 0.052 |
| 5. oxycodone (747) | 262 | 10.15% | - | 224 | 7.62% | 0.001* | 193 | 6.58% | <0.001* | 68 | 4.21% | <0.001* |
| 6. crack (749) | 152 | 5.89% | - | 178 | 6.06% | 0.82 | 228 | 7.78% | 0.007* | 191 | 11.83% | <0.001* |
| 7. methamphetamine (453) | 93 | 3.60% | - | 105 | 3.57% | 1 | 176 | 6.00% | <0.001* | 79 | 4.89% | 0.05* |
| 8. alprazolam (297) | 59 | 2.29% | - | 78 | 2.65% | 0.39 | 82 | 2.80% | 0.23 | 78 | 4.83% | <0.001* |
| 9. clonazepam (275) | 90 | 3.49% | - | 77 | 2.62% | 0.07 | 73 | 2.49% | 0.03* | 35 | 2.17% | 0.02* |
| 10. amphetamine (205) | 54 | 2.09% | - | 68 | 2.31% | 0.58 | 54 | 1.84% | 0.56 | 29 | 1.80% | 0.57 |
| 11. hydrocodone (227) | 98 | 3.80% | - | 63 | 2.14% | 0.0003* | 53 | 1.81% | <0.001* | 13 | 0.81% | <0.001* |
| 12. marijuana (235) | 80 | 3.10% | - | 83 | 2.83% | 0.58 | 44 | 1.50% | <0.001* | 28 | 1.73% | 0.007* |
| 13. fentanyl (212) | 0 | 0.00% | - | 24 | 0.82% | <0.001* | 78 | 2.66% | <0.001* | 110 | 6.82% | <0.001* |
| 14. α-PVP/bath salts (160) | 85 | 3.29% | - | 52 | 1.77% | 0.0003* | 21 | 0.72% | <0.001* | 2 | 0.12% | <0.001* |
| 15. gabapentin (148) | 18 | 0.70% | - | 63 | 2.14% | <0.001* | 41 | 1.40% | 0.01* | 26 | 1.61% | 0.007* |
| 16. methylphenidate (111) | 35 | 1.36% | - | 36 | 1.23% | 0.72 | 20 | 0.68% | 0.01* | 20 | 1.24% | 0.78 |
| 17. methadone (82) | 35 | 1.36% | - | 22 | 0.75% | 0.03* | 15 | 0.51% | 0.001* | 10 | 0.62% | 0.03* |

**Supplemental Table 3.** Charges reported to the Maine Diversion Alert Program from 2014-2017 by drug and offense category. alpha-pyrrolidinopentiophenone: α-PVP; Operating Under the Influence: OUI. ^a^chi-square with Yates’ Correction determined *p* values by comparing each offense per drug relative to all other offenses and drugs.

| Drug (N) |  | Offense |  |  |  |  |  |  |
| --- | --- | --- | --- | --- | --- | --- | --- | --- |
|  |  | Possession | Trafficking | Distribution | Possession/Intent to Distribute | Manufacturing | OUI | Other |
| 1. heroin (2,203) | N | 1,012 | 870 | 126 | 115 | 17 | 6 | 57 |
|  | % | 45.94 | 39.49 | 5.72 | 5.22 | 0.77 | 0.27 | 2.59 |
|  | p | <0.001* | <0.001* | <0.001* | <0.001* | 0.69 | <0.001* | <0.001* |
| 2. cocaine (945) | N | 552 | 186 | 90 | 70 | 2 | 1 | 44 |
|  | % | 58.41 | 19.68 | 9.52 | 7.41 | 0.21 | 0.11 | 4.66 |
|  | p | 0.31 | 0.0003* | <0.001* | <0.001* | 0.04* | 0.001* | 0.1 |
| 3. unspecified drug (906) | N | 612 | 176 | 0 | 0 | 0 | 19 | 99 |
|  | % | 67.55 | 19.43 | 0 | 0 | 0 | 2.1 | 10.93 |
|  | p | <0.001* | 0.0002* | <0.001* | <0.001* | 0.006* | 0.03* | <0.001* |
| 4. buprenorphine (812) | N | 678 | 98 | 0 | 1 | 0 | 5 | 30 |
|  | % | 83.5 | 12.07 | 0 | 0.12 | 0 | 0.62 | 3.69 |
|  | p | <0.001* | <0.001* | <0.001* | <0.001* | 0.001* | 0.12 | 0.007* |
| 5. crack (749) | N | 281 | 327 | 71 | 46 | 3 | 1 | 20 |
|  | % | 37.52 | 43.66 | 9.48 | 6.14 | 0.4 | 0.13 | 2.67 |
|  | p | <0.001* | <0.001* | <0.001* | <0.001* | 0.22 | 0.007* | <0.001* |
| 6. oxycodone (747) | N | 360 | 236 | 38 | 45 | 0 | 4 | 64 |
|  | % | 48.19 | 31.59 | 5.09 | 6.02 | 0 | 0.54 | 8.57 |
|  | p | <0.001* | <0.001* | 0.11 | 0.0002* | 0.01* | 0.09 | 0.002* |
| 7. methamphetamine (453) | N | 158 | 189 | 4 | 8 | 60 | 4 | 30 |
|  | % | 34.88 | 41.72 | 0.88 | 1.77 | 13.25 | 0.88 | 6.62 |
|  | p | <0.001* | <0.001* | 0.001* | >0.05 | <0.001* | 0.6 | 0.57 |
| 8. alprazolam (297) | N | 254 | 19 | 1 | 1 | 0 | 10 | 12 |
|  | % | 85.52 | 6.4 | 0.34 | 0.34 | 0 | 3.37 | 4.04 |
|  | p | <0.001* | <0.001* | 0.002* | 0.004* | 0.19 | 0.002* | 0.21 |
| 9. clonazepam (275) | N | 252 | 10 | 0 | 0 | 0 | 8 | 5 |
|  | % | 91.64 | 3.64 | 0 | 0 | 0 | 2.91 | 1.82 |
|  | p | <0.001* | <0.001* | 0.001* | 0.002* | 0.22 | 0.03* | 0.005* |
| 10. marijuana (235) | N | 55 | 98 | 3 | 2 | 2 | 15 | 60 |
|  | % | 23.4 | 41.7 | 1.28 | 0.85 | 0.85 | 6.38 | 25.53 |
|  | p | <0.001* | <0.001* | >0.05 | 0.04* | 0.98 | <0.001* | <0.001* |
| 11. hydrocodone (227) | N | 175 | 31 | 0 | 0 | 0 | 1 | 20 |
|  | % | 77.09 | 13.66 | 0 | 0 | 0 | 0.44 | 8.81 |
|  | p | <0.001* | 0.0002* | 0.004* | 0.006* | 0.29 | 0.41 | 0.08 |
| 12. fentanyl (212) | N | 67 | 37 | 43 | 48 | 0 | 2 | 15 |
|  | % | 31.6 | 17.45 | 20.28 | 22.64 | 0 | 0.94 | 7.08 |
|  | p | <0.001* | 0.02* | <0.001* | <0.001* | 0.32 | 0.91 | 0.55 |
| 13. amphetamine (205) | N | 176 | 13 | 0 | 0 | 0 | 4 | 12 |
|  | % | 85.85 | 6.34 | 0 | 0 | 0 | 1.95 | 5.85 |
|  | p | <0.001* | <0.001* | 0.006* | 0.01* | 0.33 | 0.56 | 0.98 |
| 14. α-PVP/bath salts (160) | N | 70 | 51 | 14 | 9 | 1 | 2 | 13 |
|  | % | 43.75 | 31.86 | 8.75 | 5.63 | 0.63 | 1.25 | 8.13 |
|  | p | <0.001* | 0.04* | 0.003* | 0.21 | 0.74 | 0.99 | 0.3 |
| 15. gabapentin (148) | N | 137 | 3 | 0 | 0 | 0 | 1 | 7 |
|  | % | 92.57 | 2.03 | 0 | 0 | 0 | 0.68 | 4.73 |
|  | p | <0.001* | <0.001* | 0.02* | 0.03* | 0.49 | 0.79 | 0.67 |
| 16. methylphenidate (111) | N | 85 | 15 | 0 | 0 | 0 | 0 | 11 |
|  | % | 76.58 | 13.51 | 0 | 0 | 0 | 0 | 9.91 |
|  | p | 0.0005* | 0.009* | 0.06 | 0.08 | 0.64 | 0.44 | 0.11 |
| 17. methadone (82) | N | 67 | 6 | 0 | 0 | 0 | 4 | 5 |
|  | % | 81.71 | 7.32 | 0 | 0 | 0 | 4.88 | 6.1 |
|  | p | <0.001* | 0.0005* | 0.12 | 0.15 | 0.8 | 0.01* | 0.94 |

**Supplemental Table 4.** Arrests by drug class and sex reported to the Maine Diversion Alert Program from 2014-2017. ^a^Fisher’s Exact Test *p* values evaluated relative to all other classes. *statistically significant

| **Class** | **Female (3,179)** |  | **Male (6,885)** |  | ***p*<sup>a</sup>** |
| --- | --- | --- | --- | --- | --- |
| opioids (4,456) | 1,404 | 44.16% | 3,052 | 44.33% | 0.90 |
| sedatives (773) | 295 | 9.28% | 478 | 6.94% | 0.015 |
| stimulants (2,535) | 751 | 23.62% | 1784 | 25.91% | 0.001* |
| hallucinogens /<br>other (597) | 139 | 4.37% | 458 | 6.65% | 0.001* |
| miscellaneous<br>pharmaceutical<br>(692) | 274 | 8.62% | 418 | 6.07% | 0.001* |
| other (1,011) | 316 | 9.94% | 695 | 10.09% | 0.84 |

**Supplemental Table 5.** Arrests by federal schedule and sex reported to the Maine Diversion Alert Program from 2014-2017. N is in parentheses. Rx: prescription but non-controlled. N/A: not applicable or unknown. ^a^Fisher’s Exact Test *p* values compared sex relative to all other Schedules.

| Schedule | Female (3,179) |  | Male (6,885) |  | *p*<sup>a</sup> |
| --- | --- | --- | --- | --- | --- |
| I (2,807) | 815 | 25.64% | 1,992 | 28.93% | <.001* |
| II (3,848) | 1,193 | 37.53% | 2,655 | 38.56% | 0.33 |
| III (834) | 250 | 7.86% | 584 | 8.48% | 0.31 |
| IV (792) | 307 | 9.66% | 485 | 7.04% | <.001* |
| V (12) | 4 | 0.13% | 8 | 0.12% | 0.90 |
| Rx (629) | 253 | 7.96% | 376 | 5.46% | <.001* |
| N/A (1,142) | 357 | 11.23% | 785 | 11.40% | .83 |

**Supplemental Table 6.** Drug charges reported to the Maine Diversion Alert Program from 2014-2017 by drug class, age, and sex. ^a^Fisher’s Exact Test *p* values comparing each age group by sex against others.

| Class | Age | Male |  | Female |  | p<sup>a</sup> |
| --- | --- | --- | --- | --- | --- | --- |
|  |  | N | % | N | % |  |
| **opioids**<br>(N = 4,456) | 18-29 | 1,104 | 68.49 | 592 | 34.91 | 0.0002* |
|  | 30-39 | 1,116 | 69.97 | 502 | 31.03 | 0.62 |
|  | 40-49 | 406 | 69.58 | 186 | 31.42 | 1 |
|  | 50-59 | 193 | 73.11 | 71 | 26.89 | 0.1 |
|  | 60+ | 39 | 76.47 | 12 | 23.53 | 0.29 |
|  | Unknown | 194 | 82.55 | 41 | 17.45 | <0.001* |
|  | Total | 3,052 | 68.49 | 1,404 | 31.51 | 0.9 |
| **stimulants**<br>(N = 2,535) | 18-29 | 724 | 70.16 | 308 | 29.84 | 0.86 |
|  | 30-39 | 570 | 66.67 | 285 | 33.33 | 0.004* |
|  | 40-49 | 243 | 69.43 | 107 | 30.57 | 0.71 |
|  | 50-59 | 128 | 79.01 | 34 | 20.99 | 0.01* |
|  | 60+ | 27 | 84.38 | 5 | 15.63 | 0.12 |
|  | Unknown | 92 | 88.46 | 12 | 11.54 | <0.001* |
|  | Total | 1,784 | 70.37 | 751 | 29.63 | 0.03* |
| **sedatives**<br>(N = 773) | 18-29 | 190 | 61.69 | 118 | 38.31 | 1 |
|  | 30-39 | 173 | 61.13 | 110 | 38.87 | 0.76 |
|  | 40-49 | 74 | 62.18 | 45 | 37.82 | 1 |
|  | 50-59 | 32 | 61.54 | 20 | 38.46 | 1 |
|  | 60+ | 9 | 81.82 | 2 | 18.18 | 0.22 |
|  | Unknown | 0 | 0 | 0 | 0 | 1 |
|  | Total | 478 | 61.84 | 295 | 38.16 | <0.001* |
| **miscellaneous**<br>**pharmaceuticals**<br>(N = 692) | 18-29 | 173 | 58.64 | 122 | 41.36 | 0.43 |
|  | 30-39 | 136 | 59.13 | 94 | 40.87 | 0.68 |
|  | 40-49 | 57 | 54.29 | 48 | 45.71 | 0.19 |
|  | 50-59 | 46 | 85.19 | 8 | 14.81 | <0.001* |
|  | 60+ | 6 | 75 | 2 | 25 | 0.49 |
|  | Unknown | 0 | 0 | 0 | 0 | 1 |
|  | Total | 418 | 60.4 | 274 | 39.6 | <0.001* |
| **hallucinogens/other**<br>(N = 597) | 18-29 | 199 | 73.16 | 73 | 26.84 | 0.07 |
|  | 30-39 | 133 | 78.7 | 36 | 21.3 | 0.52 |
|  | 40-49 | 51 | 69.92 | 23 | 31.08 | 0.11 |
|  | 50-59 | 47 | 90.38 | 5 | 9.62 | 0.02* |
|  | 60+ | 16 | 88.89 | 2 | 11.11 | 0.27 |
|  | Unknown | 12 | 100 | 0 | 0 | 0.08 |
|  | Total | 458 | 76.72 | 139 | 23.28 | <0.001* |
| **other (N = 1,011)** | 18-29 | 316 | 71.98 | 123 | 28.02 | 0.06 |
|  | 30-39 | 232 | 63.91 | 131 | 36.09 | 0.05* |
|  | 40-49 | 93 | 69.92 | 40 | 30.08 | 0.84 |
|  | 50-59 | 37 | 66.07 | 19 | 33.93 | 0.66 |
|  | 60+ | 15 | 88.24 | 2 | 11.76 | 0.11 |
|  | Unknown | 2 | 66.67 | 1 | 33.33 | 1 |
|  | Total | 695 | 68.74 | 316 | 31.26 | 0.84 |

**Supplemental Table 7.** Percentage of charges by drug and age (excludes N = 246 arrests and N = 354 substances where age was not listed) reported to the Maine Diversion Alert Program from 2014-2017. Numbers of arrests for each drug or age category are listed in parentheses. ^a^Fisher’s Exact Test *p* values relative to age 18-29.

| Drug (N) |  | Age |  |  |  |  | No age provided (354) |
| --- | --- | --- | --- | --- | --- | --- | --- |
|  |  | 18-29 (4,042) | 30-39 (3,518) | 40-49 (1373) | 50-59 (640) | 60+ (137) |  |
| 1. heroin (2,203) | N | 903 | 774 | 270 | 101 | 9 | 146 |
|  | % | 40.99% | 35.13% | 12.26% | 4.58% | 0.41% | 6.63% |
|  | *p*<sup>a</sup> |  | 0.7392 | 0.0406* | 0.0001* | <0.0001* | <0.0001* |
| 2. cocaine (945) | N | 372 | 319 | 120 | 62 | 18 | 54 |
|  | % | 39.37% | 33.76% | 12.70% | 6.56% | 1.90% | 5.71% |
|  | *p*<sup>a</sup> |  | 0.8416 | 0.6252 | 0.7137 | 0.1333 | 0.0005* |
| 3. unspecified drug (906) | N | 394 | 331 | 116 | 47 | 15 | 3 |
|  | % | 43.49% | 36.53% | 12.80% | 5.19% | 1.66% | 0.33% |
|  | *p*<sup>a</sup> |  | 0.6385 | 0.1645 | 0.058 | 0.6599 | 0.0001* |
| 4. buprenorphine (812) | N | 324 | 346 | 100 | 37 | 5 | 0 |
|  | % | 39.90% | 42.61% | 12.32% | 4.56% | 0.62% | 0.00% |
|  | *p*<sup>a</sup> |  | 0.0058* | 0.4157 | 0.0552 | 0.0736 | <0.0001* |
| 5. crack (749) | N | 308 | 238 | 110 | 44 | 7 | 42 |
|  | % | 41.12% | 31.78% | 14.69% | 5.87% | 0.93% | 5.61% |
|  | *p*<sup>a</sup> |  | 0.1543 | 0.6399 | 0.5722 | 0.3256 | 0.0074* |
| 6. oxycodone (747) | N | 254 | 252 | 100 | 71 | 18 | 52 |
|  | % | 34.00% | 33.73% | 13.39% | 9.50% | 2.41% | 6.96% |
|  | *p*<sup>a</sup> |  | 0.1282 | 0.2062 | 0.0001* | 0.0039* | <0.0001* |
| 7. methamphetamine (453) | N | 183 | 169 | 61 | 29 | 4 | 7 |
|  | % | 40.40% | 37.31% | 13.47% | 6.40% | 0.88% | 1.55% |
|  | *p*<sup>a</sup> |  | 0.5845 | 0.9401 | 1 | 0.5263 | 0.02* |
| 8. alprazolam (297) | N | 130 | 110 | 34 | 19 | 4 | 0 |
|  | % | 43.77% | 37.04% | 11.45% | 6.40% | 1.35% | 0.00% |
|  | *p*<sup>a</sup> |  | 0.8438 | 0.1248 | 0.8094 | 1 | <0.0001* |
| 9. clonazepam (275) | N | 96 | 104 | 54 | 20 | 1 | 0 |
|  | % | 34.91% | 37.82% | 19.64% | 7.27% | 0.36% | 0.00% |
|  | *p*<sup>a</sup> |  | 0.1312 | 0.0031 | 0.2722 | 0.3773 | 0.0005* |
| 10. marijuana (235) | N | 105 | 64 | 31 | 24 | 9 | 2 |
|  | % | 44.68% | 27.23% | 13.19% | 10.21% | 3.83% | 0.85% |
|  | *p*<sup>a</sup> |  | 0.0236* | 0.5495 | 0.1172 | 0.0118* | 0.011* |
| 11. hydrocodone (227) | N | 72 | 68 | 50 | 25 | 11 | 1 |
|  | % | 31.72% | 29.96% | 22.03% | 11.01% | 4.85% | 0.44% |
|  | *p*<sup>a</sup> |  | 0.6692 | <0.0001* | 0.0014* | <0.0001* | 0.0282* |
| 12. fentanyl (212) | N | 67 | 77 | 27 | 12 | 1 | 28 |
|  | % | 31.60% | 36.32% | 12.74% | 5.66% | 0.47% | 13.21% |
|  | *p*<sup>a</sup> |  | 0.1087 | 0.473 | 0.6232 | 0.7268 | <0.0001* |
| Total (10,064) | N | 4,042 | 3,518 | 1,373 | 640 | 137 | 354 |
|  | % | 40.16% | 34.96% | 13.64% | 6.36% | 1.36% | 3.52% |

**Supplemental Table 8.** Prevalence of drug charges by age and year reported to the Maine Diversion Alert Program from 2014-2017. ^a^Fisher’s Exact Test p values relative to 2014.

| Age | Years |  |  |  |  |  |  |  |  |  |  |  |
| --- | --- | --- | --- | --- | --- | --- | --- | --- | --- | --- | --- | --- |
|  | 2014 |  |  | 2015 |  |  | 2016 |  |  | 2017 |  |  |
|  | N | % | *p* | N | % | *p*<sup>a</sup> | N | % | *p*<sup>a</sup> | N | % | *p*<sup>a</sup> |
| 18-29 (3,365) | 974 | 45.22% | - | 997 | 41.58% | 0.01* | 920 | 38.83% | <0.01* | 474 | 37.26% | <0.01* |
| 30-39 (2,823) | 715 | 33.19% | - | 777 | 32.40% | 0.59 | 836 | 35.29% | 0.14 | 495 | 38.92% | 0.0008* |
| 40-49 (1,135) | 293 | 13.60% | - | 354 | 14.76% | 0.27 | 320 | 13.51% | 0.93 | 168 | 13.21% | 0.76 |
| 50-59 (507) | 121 | 5.62% | - | 153 | 6.38% | 0.29 | 148 | 6.25% | 0.38 | 85 | 6.68% | 0.21 |
| 60+ (117) | 28 | 1.30% | - | 32 | 1.33% | 1 | 34 | 1.44% | 0.7 | 23 | 1.81% | 0.25 |
| Not specified (246) | 23 | 1.07% | - | 85 | 3.54% | <0.01* | 111 | 4.69% | <0.01* | 27 | 2.12% | 0.02* |
| Total (8,193) | 2,154 | 100.00% | - | 2,398 | 100.00% | - | 2,369 | 100.00% | - | 1,272 | 100.00% | - |

**Supplemental Table 9.**
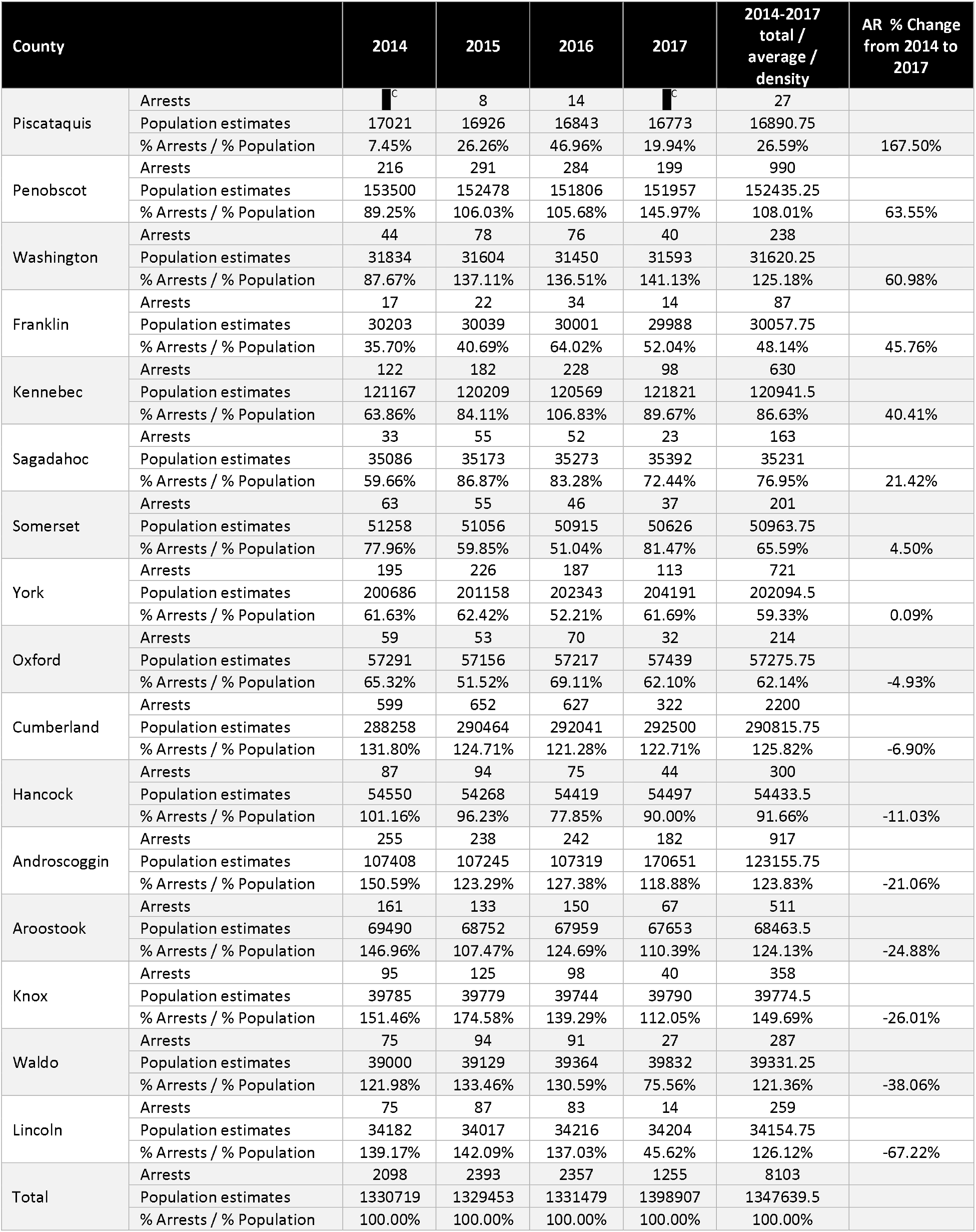
Arrests by county, ranked by the percent change in Arrest Ratio (AR), as reported to the Maine Diversion Alert Program from 2014-2017. AR is the percent arrests divided by percent population. ^C^Values ≤ 5 obscured to protect arrestee confidentiality.

| County |  | 2014 | 2015 | 2016 | 2017 | 2014-2017<br>total /<br>average /<br>density | AR % Change<br>from 2014 to<br>2017 |
| --- | --- | --- | --- | --- | --- | --- | --- |
| Piscataquis | Arrests | <sup>C</sup> | 8 | 14 | <sup>C</sup> | 27 |  |
|  | Population estimates | 17021 | 16926 | 16843 | 16773 | 16890.75 |  |
|  | % Arrests / % Population | 7.45% | 26.26% | 46.96% | 19.94% | 26.59% | 167.50% |
| Penobscot | Arrests | 216 | 291 | 284 | 199 | 990 |  |
|  | Population estimates | 153500 | 152478 | 151806 | 151957 | 152435.25 |  |
|  | % Arrests / % Population | 89.25% | 106.03% | 105.68% | 145.97% | 108.01% | 63.55% |
| Washington | Arrests | 44 | 78 | 76 | 40 | 238 |  |
|  | Population estimates | 31834 | 31604 | 31450 | 31593 | 31620.25 |  |
|  | % Arrests / % Population | 87.67% | 137.11% | 136.51% | 141.13% | 125.18% | 60.98% |
| Franklin | Arrests | 17 | 22 | 34 | 14 | 87 |  |
|  | Population estimates | 30203 | 30039 | 30001 | 29988 | 30057.75 |  |
|  | % Arrests / % Population | 35.70% | 40.69% | 64.02% | 52.04% | 48.14% | 45.76% |
| Kennebec | Arrests | 122 | 182 | 228 | 98 | 630 |  |
|  | Population estimates | 121167 | 120209 | 120569 | 121821 | 120941.5 |  |
|  | % Arrests / % Population | 63.86% | 84.11% | 106.83% | 89.67% | 86.63% | 40.41% |
| Sagadahoc | Arrests | 33 | 55 | 52 | 23 | 163 |  |
|  | Population estimates | 35086 | 35173 | 35273 | 35392 | 35231 |  |
|  | % Arrests / % Population | 59.66% | 86.87% | 83.28% | 72.44% | 76.95% | 21.42% |
| Somerset | Arrests | 63 | 55 | 46 | 37 | 201 |  |
|  | Population estimates | 51258 | 51056 | 50915 | 50626 | 50963.75 |  |
|  | % Arrests / % Population | 77.96% | 59.85% | 51.04% | 81.47% | 65.59% | 4.50% |
| York | Arrests | 195 | 226 | 187 | 113 | 721 |  |
|  | Population estimates | 200686 | 201158 | 202343 | 204191 | 202094.5 |  |
|  | % Arrests / % Population | 61.63% | 62.42% | 52.21% | 61.69% | 59.33% | 0.09% |
| Oxford | Arrests | 59 | 53 | 70 | 32 | 214 |  |
|  | Population estimates | 57291 | 57156 | 57217 | 57439 | 57275.75 |  |
|  | % Arrests / % Population | 65.32% | 51.52% | 69.11% | 62.10% | 62.14% | -4.93% |
| Cumberland | Arrests | 599 | 652 | 627 | 322 | 2200 |  |
|  | Population estimates | 288258 | 290464 | 292041 | 292500 | 290815.75 |  |
|  | % Arrests / % Population | 131.80% | 124.71% | 121.28% | 122.71% | 125.82% | -6.90% |
| Hancock | Arrests | 87 | 94 | 75 | 44 | 300 |  |
|  | Population estimates | 54550 | 54268 | 54419 | 54497 | 54433.5 |  |
|  | % Arrests / % Population | 101.16% | 96.23% | 77.85% | 90.00% | 91.66% | -11.03% |
| Androscoggin | Arrests | 255 | 238 | 242 | 182 | 917 |  |
|  | Population estimates | 107408 | 107245 | 107319 | 170651 | 123155.75 |  |
|  | % Arrests / % Population | 150.59% | 123.29% | 127.38% | 118.88% | 123.83% | -21.06% |
| Aroostook | Arrests | 161 | 133 | 150 | 67 | 511 |  |
|  | Population estimates | 69490 | 68752 | 67959 | 67653 | 68463.5 |  |
|  | % Arrests / % Population | 146.96% | 107.47% | 124.69% | 110.39% | 124.13% | -24.88% |
| Knox | Arrests | 95 | 125 | 98 | 40 | 358 |  |
|  | Population estimates | 39785 | 39779 | 39744 | 39790 | 39774.5 |  |
|  | % Arrests / % Population | 151.46% | 174.58% | 139.29% | 112.05% | 149.69% | -26.01% |
| Waldo | Arrests | 75 | 94 | 91 | 27 | 287 |  |
|  | Population estimates | 39000 | 39129 | 39364 | 39832 | 39331.25 |  |
|  | % Arrests / % Population | 121.98% | 133.46% | 130.59% | 75.56% | 121.36% | -38.06% |
| Lincoln | Arrests | 75 | 87 | 83 | 14 | 259 |  |
|  | Population estimates | 34182 | 34017 | 34216 | 34204 | 34154.75 |  |
|  | % Arrests / % Population | 139.17% | 142.09% | 137.03% | 45.62% | 126.12% | -67.22% |
| Total | Arrests | 2098 | 2393 | 2357 | 1255 | 8103 |  |
|  | Population estimates | 1330719 | 1329453 | 1331479 | 1398907 | 1347639.5 |  |
|  | % Arrests / % Population | 100.00% | 100.00% | 100.00% | 100.00% | 100.00% |  |

**Supplemental Table 10.**
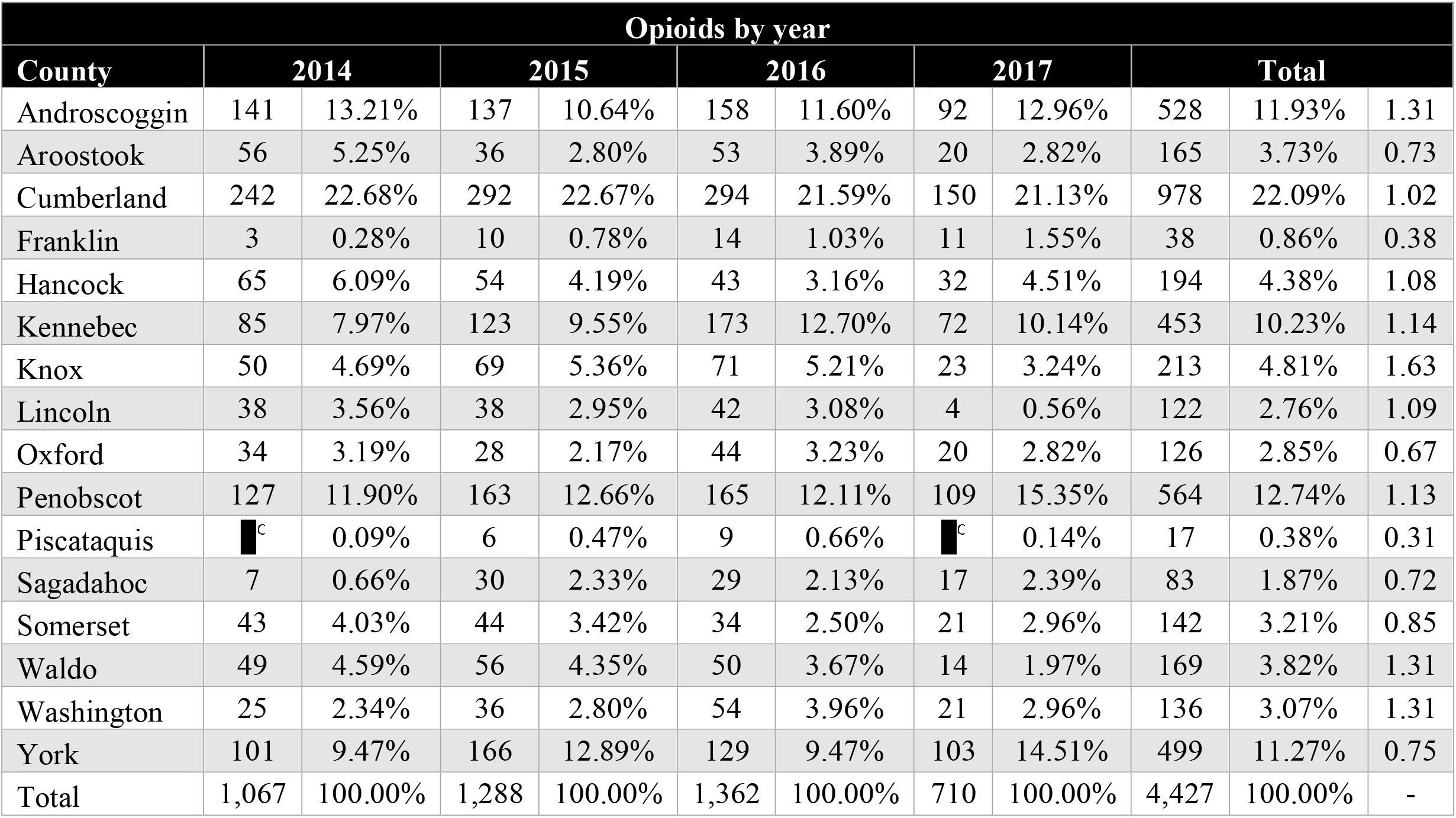
Opioid arrests by county reported to the Maine Diversion Alert Program from 2014-2017. ^C^Values ≤ 5 are obscured to protect arrestee confidentiality.

**Supplemental Table 11.**
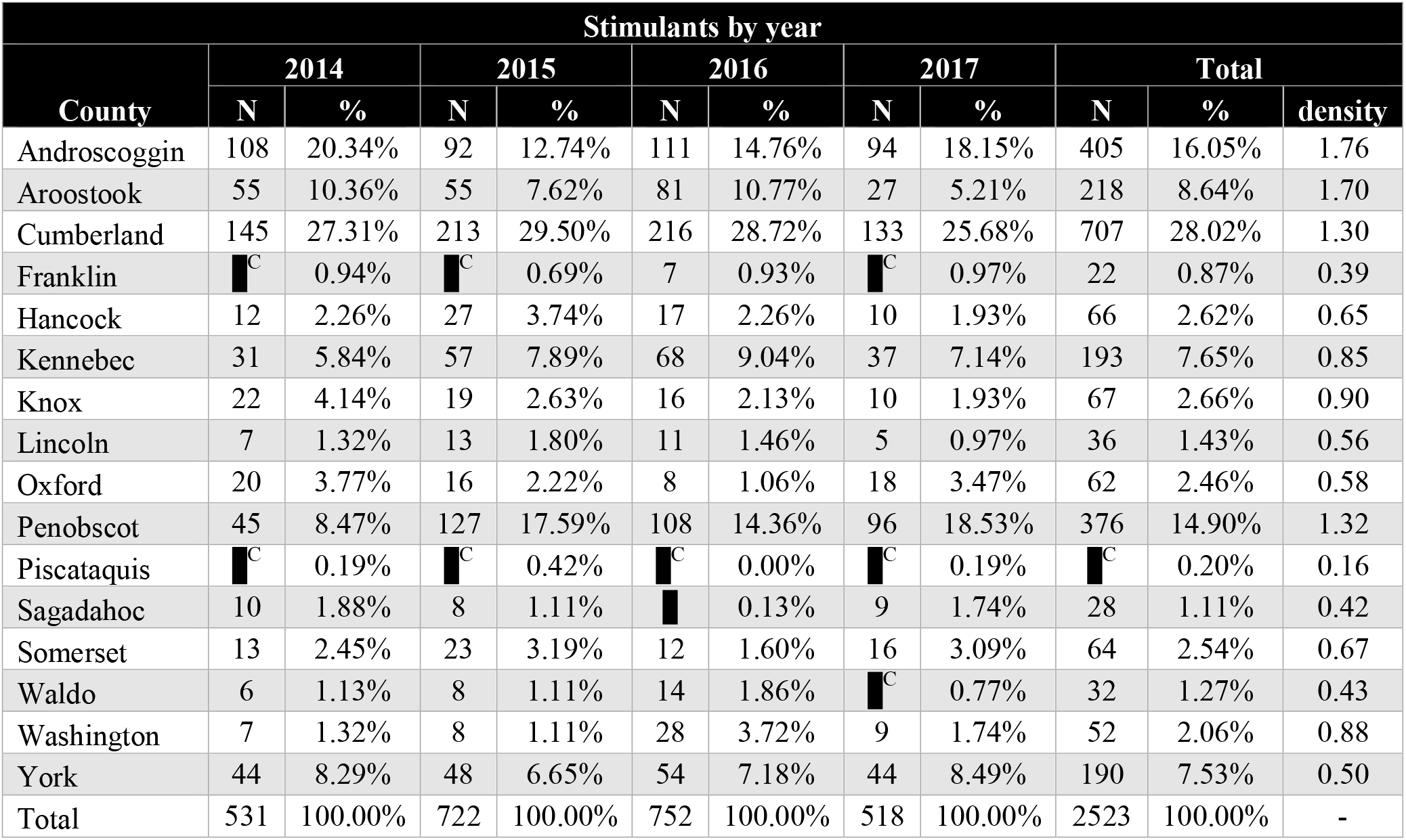
Stimulant arrests by county reported to the Maine Diversion Alert Program from 2014-2017. ^C^Values ≤ 5 obscured to protect arrestee confidentiality.

| Stimulants by year |  |  |  |  |  |  |  |  |  |  |  |
| --- | --- | --- | --- | --- | --- | --- | --- | --- | --- | --- | --- |
| County | 2014 |  | 2015 |  | 2016 |  | 2017 |  | Total |  |  |
|  | N | % | N | % | N | % | N | % | N | % | density |
| Androscoggin | 108 | 20.34% | 92 | 12.74% | 111 | 14.76% | 94 | 18.15% | 405 | 16.05% | 1.76 |
| Aroostook | 55 | 10.36% | 55 | 7.62% | 81 | 10.77% | 27 | 5.21% | 218 | 8.64% | 1.70 |
| Cumberland | 145 | 27.31% | 213 | 29.50% | 216 | 28.72% | 133 | 25.68% | 707 | 28.02% | 1.30 |
| Franklin | <sup>C</sup> | 0.94% | <sup>C</sup> | 0.69% | 7 | 0.93% | <sup>C</sup> | 0.97% | 22 | 0.87% | 0.39 |
| Hancock | 12 | 2.26% | 27 | 3.74% | 17 | 2.26% | 10 | 1.93% | 66 | 2.62% | 0.65 |
| Kennebec | 31 | 5.84% | 57 | 7.89% | 68 | 9.04% | 37 | 7.14% | 193 | 7.65% | 0.85 |
| Knox | 22 | 4.14% | 19 | 2.63% | 16 | 2.13% | 10 | 1.93% | 67 | 2.66% | 0.90 |
| Lincoln | 7 | 1.32% | 13 | 1.80% | 11 | 1.46% | 5 | 0.97% | 36 | 1.43% | 0.56 |
| Oxford | 20 | 3.77% | 16 | 2.22% | 8 | 1.06% | 18 | 3.47% | 62 | 2.46% | 0.58 |
| Penobscot | 45 | 8.47% | 127 | 17.59% | 108 | 14.36% | 96 | 18.53% | 376 | 14.90% | 1.32 |
| Piscataquis | <sup>C</sup> | 0.19% | <sup>C</sup> | 0.42% | <sup>C</sup> | 0.00% | <sup>C</sup> | 0.19% | <sup>C</sup> | 0.20% | 0.16 |
| Sagadahoc | 10 | 1.88% | 8 | 1.11% | <sup>C</sup> | 0.13% | 9 | 1.74% | 28 | 1.11% | 0.42 |
| Somerset | 13 | 2.45% | 23 | 3.19% | 12 | 1.60% | 16 | 3.09% | 64 | 2.54% | 0.67 |
| Waldo | 6 | 1.13% | 8 | 1.11% | 14 | 1.86% | <sup>C</sup> | 0.77% | 32 | 1.27% | 0.43 |
| Washington | 7 | 1.32% | 8 | 1.11% | 28 | 3.72% | 9 | 1.74% | 52 | 2.06% | 0.88 |
| York | 44 | 8.29% | 48 | 6.65% | 54 | 7.18% | 44 | 8.49% | 190 | 7.53% | 0.50 |
| Total | 531 | 100.00% | 722 | 100.00% | 752 | 100.00% | 518 | 100.00% | 2523 | 100.00% | - |

## Discussion

This study spanned four years of data involving over eight-thousand drug-related arrests and ten-thousand drug charges. This report is the largest and most thorough analysis of the Maine DAP to date [5, 11, 13, 14] and enables a comprehensive assessment of trends and demographic subgroup examination from a data source unlike any other in the US. Well-meaning regulatory and administrative barriers functioned to effectively ban US prisoners from much research [23] resulting in this IRB approved dataset being rare and valuable. The analysis of drug arrests and patterns of drug charges in this report may be used to guide public health policy and optimize resource utilization. The most unique aspect of this report is detailed reporting of the offenses involved (Figures 1-2) and examination by demographic characteristics (Figures 3-4).

Nationally and in Maine, the opioid epidemic can be visualized as three distinct waves. The first opioid wave of drug crimes and overdose deaths involving prescription drugs appears to be ebbing. For example, drug charges for prescription oxycodone and hydrocodone reported to the Maine DAP were significantly lower in 2015, 2016 and 2017 compared to 2014 (Figure 1B). Opioid prescribing has undergone sizable changes in Maine between 2012 and 2017 with more than a forty-percent reduction in fentanyl, morphine, tapentadol, meperidine, hydrocodone, and oxydocone but a 58% increase in buprenorphine [5]. Other states with opioid prescribing laws have not noticed pronounced opioid reductions [24]. The second opioid wave of drug crimes and overdose deaths involving heroin may be cresting. In Maine, drug charges for heroin peaked in 2016 and were significantly higher in 2016 and 2017 compared to 2014. This pattern in Maine of heroin drug charges is consistent with heroin overdose deaths which also peaked in 2016 and subsequently declined [5]. The third wave of illicit fentanyl use and overdose death poses an alarming and increasing threat to Maine and the US. Drug charges in Maine for fentanyl were significantly higher in 2015, 2016 and 2017 compared to 2014. Likewise, overdose deaths from illicit fentanyl continue to rise in Maine with 247 fatalities in 2017 compared to only one in 2013 [5].

The adverse impacts from excessive distribution of opioid prescription drugs may be declining in Maine in part due to recent laws that limit opioid doses and mandate use of the PDMP thereby reducing overprescribing and diversion [8]. Maine’s opioid prescribing law is unique in the US to include financial penalties for non-adherence [1]. Nonetheless, nonmedical use of opioid prescription drugs remains prevalent especially in older adults (Figure 3A). Complicating the picture, the successful reduction in availability of opioid prescriptions [5] coupled with a lack of available and timely treatment for those with opioid use disorder may be inadvertently contributing to the shift toward cheaper and more easily obtainable heroin and illicit fentanyl. The recent rise in illicit fentanyl arrests and overdose deaths in Maine reflect the dramatic escalation of the opioid epidemic. Only two milligrams of fentanyl is a potentially lethal dose [25]. Recent arrests in Maine and New Hampshire have resulted in the seizure of over 30 kilograms of illicit fentanyl. This is enough to potentially cause 15 million overdoses [26-28] which is over five-fold greater than the population of these states.

There were over eight-hundred arrests for buprenorphine making this mu partial agonist the third most common individual drug reported to the DAP and the most common prescription drug. The preponderance (84%) of buprenorphine arrests were for possession. The median street value for the buprenorphine/naloxone film among Australian persons who inject drugs was $31 for 8 mg [19]. A US online reporting system lists values in 2019 between $8-30 for 8 mg [29]. This report extends upon earlier ones [2, 11, 14] and found that buprenorphine accounted for one-tenth of arrests for individuals in their thirties. Given the increased availability of this OUD treatment in Maine [5] and nationwide [1], additional qualitative study of arrestees is needed to determine if this is the result of self-medication, non-medical use [16, 30], or some other factors.

While the current opioid crisis has deservedly garnered significant attention, stimulants remain prevalent, the use of cocaine and crack cocaine appears to have rebounded and non-medical use of sedatives such as prescription alprazolam is increasing in Maine. Drug arrests for methamphetamine were significantly higher in 2016 and 2017 than 2014 (Figure 1B). Meanwhile, drug arrests for cocaine/crack cocaine and alprazolam were significantly higher in 2017 compared to 2014 in Maine. Drug arrests involving stimulants were significantly more likely among men while drug arrests for sedatives such as alprazolam and non-controlled prescription drugs were significantly higher among women (Figure 3B). The biological or environmental hypothesis that might explain a difference in nonmedical drug use between men and women such that men are more likely to be arrested for stimulants and women are more likely to be arrested for sedatives remains to be determined. Previous exposure to similar prescription drugs has contributed to this phenomenon. Prescription stimulants are indicated for attention deficit hyperactivity disorder which is more prevalent in men whereas sedatives, specifically benzodiazepines, are indicated for anxiety disorders which are more prevalent among women [31, 32].

The increasing prevalence of drug arrests for these substances needs careful monitoring in both sexes. Although naloxone availability has expanded greatly in this state, almost one-third of overdose decedents in Maine tested positive for naloxone [5]. This could imply that either naloxone was administered too late, the dose was insufficient to overcome the mu receptor agonist (e.g. carfentanil), that other substances were involved, or that naloxone was bioavailable via injection of buprenorphine/naloxone. There is also increasing evidence that sublingual administered naloxone in buprenorphine/naloxone products is bioavailable to clinically significant degrees [33]. The arrest data indicate that increased resources for the provision of evidence based interventions [34] for cocaine/crack cocaine may be necessary in the near future.

The arrest information communicated from law-enforcement to medical professionals by the DAP complemented that provided by the state’s PDMP [10]. Future modifications and implementation of the DAP in other municipalities could include: 1) raising the profile of this type of harm-reduction tool with relevant stake-holders; 2) integration of a DAP with the PDMP so that busy health care providers do not need to register and access multiple databases; 3) mandatory data uploading to a secure server from all local, county, and tribal law enforcement agencies; 4) inclusion of arrest information about minors (≥16) or arrestee ethnicity to better characterize the needs of diverse populations; 5) differentiation of pharmaceutical versus illicit formulations (e.g. for fentanyl); 6) identification of a stable funding source; and 7) availability of updated (i.e. post-arrest) information about the analytical chemistry of drugs (i.e. there are serious concerns about the transparency, limit of detection, selectivity and specificity of the “spot” or field tests [35]) and final judicial outcomes.

Over four-fifths of benzodiazepine arrests were for possession. In contrast, possession was less common than trafficking for marijuana. Further study is needed to better understand the generalizability of the findings in Figure 2 as other states have different offense criteria. The weight disparity for cocaine versus crack cocaine continues to warrant further attention as the so-called “Fair” Sentencing Act of 2010 only attenuated the disparities for crack cocaine versus cocaine [36].

This report extends and integrates findings from prior single-year reports [5, 11, 13, 14]. Some caveats with the DAP have been discussed previously [11]. Briefly, this well powered dataset identified some findings, for example males accounted for 2.3% more stimulant arrests, which, although statistically significant, might be of limited value for health care providers. Sex differences in criminal justice system involvement, and also overdoses, may be becoming less pronounced. It would be of substantial value if DAP included information about the formulation so as to know which ones were most common among the eight-hundred buprenorphine arrests. This data, as well as that obtained by others [20], indicates that the nonmedical use potential of buprenorphine should be clearly emphasized in X-waiver trainings. State laws may limit the generalizability of select findings to other areas. For example, Maine approved medical cannabis in 1999 and recreational marijuana in 2016 which likely contributed to the low occurrence of arrests for marijuana possession. The limited ethnic diversity of Maine is another important consideration. The age ranges at the extremes (i.e. 18-29, ≥ 60) were not equivalent in terms of years covered or the number of arrestees which could have a minor impact on the results. This dataset was deidentified for investigative purposes which precluded examination of arrests among repeat offenders.

Further, arrests reflect a combination of use patterns and law enforcement priorities. This pharmacoepidemiology index could be viewed as complementing anonymous surveys, prescriptions of licit drugs, emergency room visits, and medical examiner reports.

In conclusion, this multiyear study reveals a complex and evolving nonmedical drug use patterns. Heroin and illicit fentanyl are the greatest concerns while prescription opioid nonmedical use remains a persistent problem. Recent increases in cocaine/crack cocaine, methamphetamine, and alprazolam related crimes may signal the emergence of new patterns of non-medical drug use that warrant further monitoring. The overall drug situation continues to worsen as evidenced by rising overdose deaths and further national public health policy initiatives are needed to reverse these trends.

## Data Availability

Data is available as an Excel spreadsheet.

